# Risky sexual behavior and its determinants among secondary school students in the highly HIV/AIDS burdened setting of South Ethiopia: Implications for decision makers

**DOI:** 10.1101/2023.07.05.23292257

**Authors:** Getamesay Aynalem Tesfaye, Ermias Wabeto Wana, Maranata Dawit Ambaw, Bewuket Addis Alemayehu

**Affiliations:** Public Health Department, College of Medicine and Health Sciences, Jinka University, Jinka, Ethiopia; Nursing Department, College of Medicine and Health Sciences, Jinka University, Jinka, Ethiopia

**Keywords:** Risky Sexual Behaviors, HIV/AIDS, High School Students, South Omo Zone, Ethiopia

## Abstract

**Background:** Risky sexual behavior (RSB) among youth can lead to serious social, economic, and health problems, such as unwanted pregnancy and acquired immune deficiency syndrome (AIDS). Despite a high burden of AIDS in the South Omo zone, little is known about RSB among secondary school students, and numerous studies in Ethiopia have discordant RSB findings. Therefore, this study aimed to assess the magnitude of risky sexual behavior and its determinants among secondary school students in the South Omo zone.

**Methods:** An institution-based cross-sectional study was conducted in January 2023. A multistage sampling method was used to select 538 students. A validated and structured self-administered questionnaire was used to collect data after pretesting. The results of descriptive analysis were presented by texts, tables and figures. Variables found to be p<0.20 in the bivariate logistic regression analysis were candidates for multivariate analysis. The level of statistical significance was declared at a p value less than 0.05 in multivariate analysis. Odds ratios along with their 95% confidence intervals (CIs) were used to present the results of the logistic regressions.

**Results:** The prevalence of RSB among the study participants was 25.9% (95% CI= 22.3%-29.9%). It was significantly associated with having sexually active close friends [adjusted odds ratio (AOR) = 3.09; 95% CI: 1.90-5.02], attending night clubs (AOR=2.56; 95% CI: 1.35-4.86), drinking alcohol (AOR=1.90; 95% CI: 1.10-3.29), parental neglect (AOR=2.10; 95% CI: 1.35-3.29) and HIV/AIDS-related knowledge (AOR=1.76; 95% CI: 1.12-2.77).

**Conclusions:** Risky sexual behavior among secondary school students was very high. Having sexually active friends, attending night clubs, drinking alcohol, parental neglect and HIV/AIDS-related knowledge were determinants of RSB. Strengthening reproductive and sexual health services, close communication with youth in the family, measures to lessen alcohol consumption and night club attendance, and enhancing HIV/AIDS-related knowledge among secondary school students are needed.

**Plain English summary:** Young people, who are aged 10 to 24 years, face various reproductive health problems as they transition from childhood to adulthood, including risky sexual behavior. Risky sexual behavior includes having more than one sexual partner, having first sex before 18 years, not usually using condoms during sexual intercourse, or having sex with commercial sex workers. It could lead to health problems such as unwanted pregnancy and sexually transmitted diseases.

This study was conducted to assess the amount of risky sexual behavior and related factors among secondary school students in the South Omo zone, South Ethiopia. By chance, we selected 538 students aged 15 to 24 years attending grades 9 to 12. Data were collected by using a questionnaire that selected students answered and returned. The data were processed and analyzed by a personal computer.

Among all selected students, 97% participated in the study. The current study showed that approximately one out of four students were practicing risky sexual behavior. The occurrence of risky sexual behavior was high among students with sexually active close friends, night club attendance, alcohol consumption habits, parental neglect and low HIV/AIDS-related knowledge. In conclusion, the authors recommend decision makers intensify the availability and accessibility of reproductive and sexual health services for youths. Parents should be encouraged to openly and appropriately discuss reproductive and sexual health issues with their children. Additionally, it is crucial to decrease alcohol drinking habits and night club attendance of students, in addition to increasing HIV/AIDS-related knowledge.

## Background

Young people are comprised of girls and boys whose ages range between 10 and 24 years, spanning the periods of adolescence and youth, which is also the age of most secondary school students. As young people transition from childhood to adulthood, they undergo rapid physical and psychosocial changes, and they gain new experiences, acquire new capacities and face many new situations and challenges (1). During this transition, they must be prepared with the knowledge and skills they need to make use of the opportunities and to face the challenges they will encounter in the adult world (2).

Mostly risky sexual behavior (RSB) is defined based on the behavior itself, which encompasses early sexual activity and unprotected sexual activity, the number and types of sexual partners, relationship to partner, frequency of sexual intercourse, and condom use (3, 4). The other but neglected way of RSB definition refers to the nature of the partner that includes if a partner is human immunodeficiency virus (HIV) positive, whether a partner is an intravenous drug user, or if a partner is nonexclusive. RSB among youth can lead to serious social, economic, and health problems such as sexual violence, unwanted pregnancy, ectopic pregnancy, cervical cancer, infertility and sexually transmitted diseases, including HIV infection (5–7).

Despite the effort of the World Health Organization to end the AIDS (Acquired Immunodeficiency Disease) epidemic as a public health threat by 2030, new HIV infections and AIDS-related deaths remain unacceptably high (8, 9). In 2020, youths contributed 44% of new HIV infections in sub-Saharan Africa despite comprising only 20% of the population (9). In Ethiopia, 0.2% of youths aged 15-24 are HIV-positive (10).

Ethiopia has developed strategies to meet the sexual and reproductive health demands of young people in the country and to end AIDS as a public health threat in 2030 (10, 11). However, the country has many RSB challenges among young people who have yet to be addressed. For instance, in 2016, 12.56% and almost 5% of youth people ever chewed khat and drank alcohol every day, respectively, and one in four women and 2% of men had first sexual intercourse before their fifteenth birthday (12). Additionally, numerous other studies in Ethiopia have underscored the RSB problem among young people (13–15).

Despite a high burden of HIV infections, little is known about the magnitude of RSB among secondary school students in the South Omo zone, South Ethiopia. Furthermore, numerous studies in Ethiopia on RSB had discordant findings (16–18). Therefore, this study was designed to determine the prevalence of RSB and its determinants among secondary school students in the South Omo zone. Therefore, the study will have implications for RSB and its determinants for decision makers to plan and manage RSB problems, especially HIV infections, among youths in the study area.

## Methods

### Study design and period

An institution-based cross-sectional study was conducted from January 1 to January 31, 2023.

### Study setting

The study was conducted in the South Omo zone, which is located 767 kilometers southwest of Addis Ababa, the capital city of Ethiopia. The South Omo zone forms part of the Ethiopian region of SNNPR and is based in Jinka as its administrative town (19). It is one of Ethiopia’s socially most diverse zones, containing a minimum of 12 different ethnic groups and possibly as many as 21. There are 820,480 people in nine districts of the South Omo zone who live as pastoralists and/or agrarians. In the South Omo zone, there are 21,848 students in 35 secondary schools comprising 11,651 male students and 10,197 female students.

### Source and study population

The source population for this study was all secondary school students in the South Omo zone, whereas the study population was all selected secondary school students in the South Omo zone. The study unit was a randomly selected student from the selected secondary school students in the South Omo zone.

### Eligibility criteria

All regular students aged 15 to 24 years who were registered in a selected secondary school were included in the study. All secondary school students who were unable to respond due to sickness or those students who were not attending regular classes were excluded from the study due to data collection difficulties.

### Sample size determination

The sample size for this study was determined using a single population proportion formula. Considering the following assumptions: 95% confidence level, 5% marginal error and 30.5% prevalence of RSB from a study in Mizan Aman (13), the formula yields a sample size of 326. After adding 10% for the nonresponse rate and with a design effect of 1.5, the required final sample size became 538.

### Sampling procedure

A multistage sampling method was employed to select study participants. Hence, initially, three districts were selected randomly from all districts in the South Omo zone. Then, the calculated sample size was allocated proportionally to the three districts based on their number of secondary school students. Using the school register list as a sampling frame, a systematic random sampling technique was applied to select students from secondary school in each district.

### Study variables

Risky sexual behavior is the outcome variable of the study. On the other hand, the independent variables are sociodemographic variables such as sex, age, grade level, religiosity, residence, living out of family, educational level of parents, occupation of parents, pocket money, and discussion on sexual and reproductive issues. HIV/AIDS knowledge, parental neglect, social support, and personal behaviors such as drinking alcohol, chewing khat, watching pornography, and attending night clubs are also independent variables of the study.

### Operational definitions and variable measurements

Risky sexual behavior was defined as a student practicing at least one of the following behaviors: multiple sexual partners (having more than one sexual partner until the survey), early initiation of sex (first sex at <18 years), inconsistent use of condom (inconsistent/fail to use condom at least once during sexual intercourse until the survey), and sex with commercial sex workers at least once until the survey) (20). Adapted from the Anne Murphy study, parental neglect was measured by an adverse childhood experience questionnaire asking about childhood abuse (emotional, physical and sexual), childhood neglect (emotional and physical) and household dysfunction (mother treated violently, parental separation or divorce, mental illness in household, household substance abuse, incarcerated household member) (21). Consequently, students with adverse childhood experience scores above the mean were proclaimed to be victims of parental neglect. Social support was assessed by the Oslo Social Support Scale, which has three items. The sum of the scores will be categorized into three social support levels: poor (3–8), moderate (9–11), and strong (12–14)(22). Respondents were declared to have good knowledge about HIV/AIDS when they answered HIV/AIDS-related questions correctly more than the mean value (18).

### Data collection

A validated structured self-administered questionnaire adapted from the Youth Risk Behavior Survey questionnaire was used to collect data (23). The questionnaire comprised questions about sociodemographic factors, personal behaviors, parental neglect, social support, and so on. It was prepared in the English language and translated into the Amharic language and then back to English by fluent speakers to check meaning consistency.

### Data quality control

All research assistants were given two days of intensive training on the purpose of the study, ethical issues and how to facilitate the data collection. The questionnaire was pretested, two weeks prior to the actual data collection period, on regular government secondary school students with a size of 5% of the final sample size in nonselected districts. After pretesting the questionnaire, the reliability of the tool was assessed, and necessary modifications were made. The research investigators and the supervisors checked the completeness of each questionnaire. Two data clerks entered the data, and the consistency was cross checked. Multivariable logistic regression analysis was run to control confounding factors.

### Data processing and analysis

After coding and cleaning, the data were entered into and analyzed by SPSS v26. Descriptive statistical analysis was used to describe the sociodemographic and sexual characteristics of the participants. Then, the information was presented using summary measures, tables, figures and texts. Both bivariate and multivariate logistic regression analyses were employed to determine the association between the output variable and independent variables. The multicollinearity effect of independent variables was checked, and variables with variance inflation factors (VIFs) greater than 5 were dropped from bivariate analysis. All covariates that were significant at p value < 0.20 in bivariate analysis were considered for further multivariate analysis. The level of statistical significance was declared at a p value less than 0.05 in multivariate analysis, and the fitness of the multivariable logistic regression model was assured by the Hosmer-Lemeshow goodness of fit test and classification table. Crude and adjusted odds ratios along with their 95% confidence intervals were used to present the results of logistic regression.

## Results

### Sociodemographic characteristics

Among 538 selected students, 522 participated in the study, yielding a response rate of 97%. The median age of the students was 17.44 years, with a range of 9 years. Approximately 58.4% and 81% of students’ mothers and fathers were able to read and write, respectively. The monthly family income was less than 5,000 Ethiopian Birr for most (58%) students. Almost half (48.5%) of the participants discussed sexual and reproductive health issues with their parents, while one-third (32.2%) of them felt peer pressure about sexual and reproductive health issues (Table 1).

**Table 1:** Sociodemographic characteristics of secondary school students in the South Omo zone, Southwest Ethiopia, 2023 (N= 522).

| Variables | Category | Frequency |  |
| --- | --- | --- | --- |
|  |  | Number<br>(n) | Percent<br>(%) |
| Age group | 15–19 years old | 471 | 90.2 |
|  | 20–24 years old | 51 | 9.8 |
| Sex | Male | 274 | 52.5 |
|  | Female | 248 | 47.5 |
| Grade | 9th grade | 138 | 26.4 |
|  | 10th grade | 118 | 22.6 |
|  | 11th grade | 135 | 25.9 |
|  | 12th grade | 131 | 25.1 |
| Place of primary school | Urban | 306 | 58.6 |
|  | Rural | 216 | 41.4 |
| **Living with parents** | Yes | 415 | 79.5 |
|  | No | 107 | 20.5 |
| **Parental monitoring** | Yes | 385 | 73.8 |
|  | No | 137 | 26.2 |
| **Pocket money** | Yes | 296 | 56.7 |
|  | No | 226 | 43.3 |
| **Visit of religious institution** | Never | 29 | 5.6 |
|  | Sometimes | 168 | 32.2 |
|  | Often | 325 | 62.3 |
| **Close friend experienced sex** | Yes | 110 | 21.1 |
|  | No | 412 | 78.9 |
| **Social support** | Poor | 229 | 43.9 |
|  | Moderate | 200 | 38.3 |
|  | Strong | 93 | 17.8 |

### Sexual behavior and drug use

The prevalence of risky sexual behavior among secondary school students in the South Omo zone is 25.9% (95% CI: 22.3-29.9). One-fourth (23.2%) of respondents had ever had sexual intercourse, while the mean age of sexual debut was 15.84 years, with a standard deviation of 2.1 years. Of all respondents who ever had sexual intercourse, 57% and 87.6% of them had sexual intercourse in the past 1 year and did not always use condoms, respectively. The main reasons for not using condoms were trusting sexual partners (54.7%), unavailability of condoms (13.2%), decrease in sexual pleasure (20.8%), refusal by sexual partners (7.2%) and others (4.3%) (Figure 1) (Table 2).

**Figure 1:** Reasons to start sexual intercourse among secondary school students in the South Omo zone (N= 121).

**Table 2:**
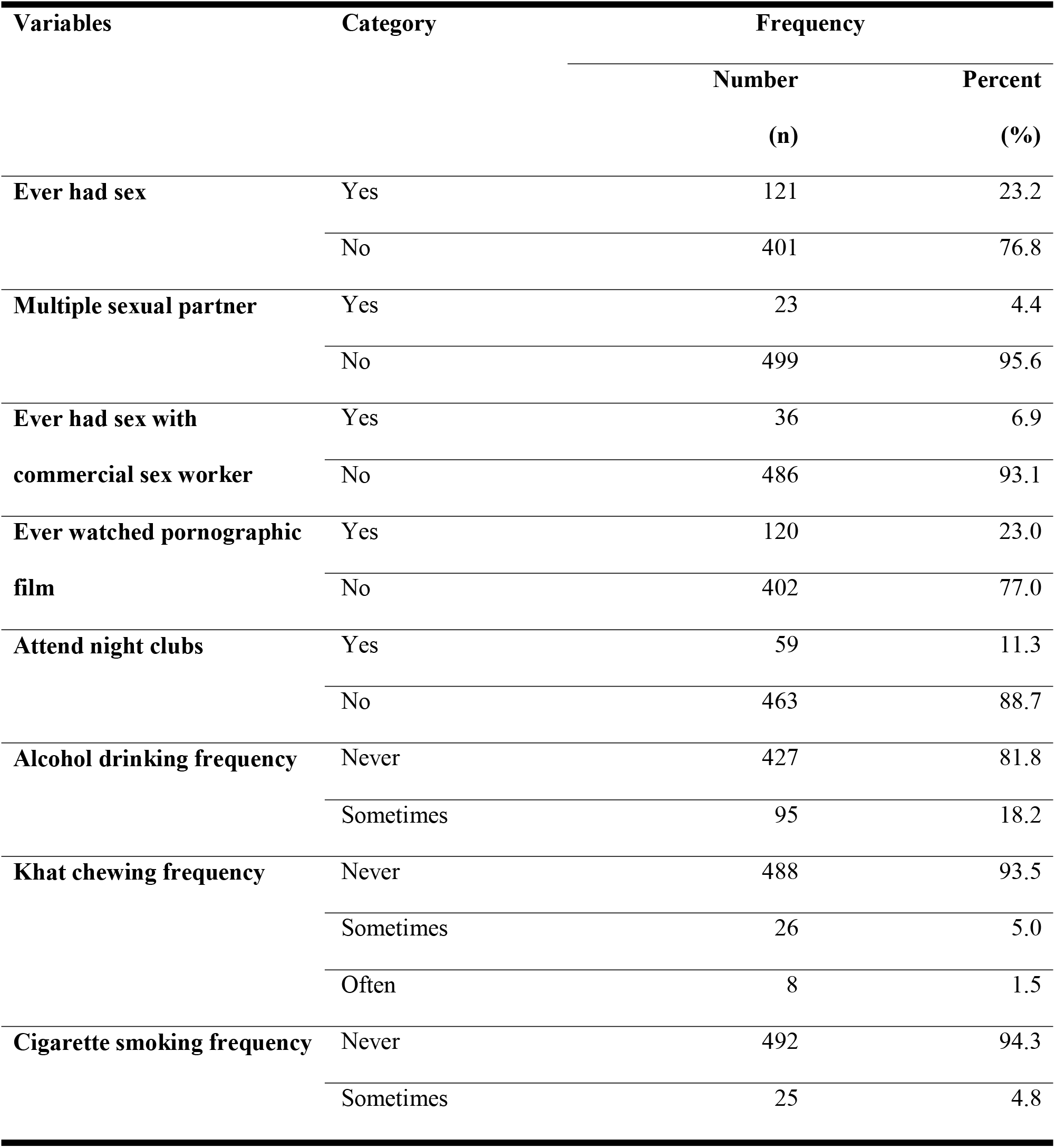
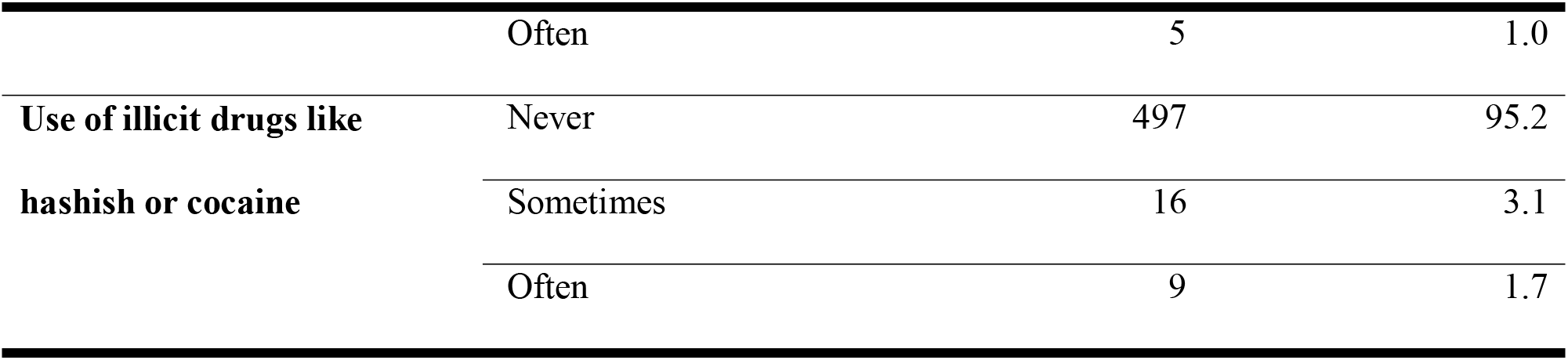
Sexual behavior and drug use of respondents (N= 522).

### Determinants of risky sexual behavior

During multivariate logistic regression, close friends’ sexual experience, night club attendance, alcohol drinking habits, parental neglect, and HIV/AIDS knowledge were identified as determinants of risky sexual behavior (Table 3).

**Table 3:** Determinants of risky sexual behavior among secondary school students in the South Omo zone, 2023 (N= 522).

| Variables | Risky sexual behavior |  | COR | AOR |
| --- | --- | --- | --- | --- |
|  | No (387) | Yes (135) | (95% CI) | (95% CI) |
| **Close friend experienced sex** |  |  |  |  |
| No | 331 | 81 | 1 | 1 |
| Yes | 56 | 54 | 3.94(2.52-6.15)** | 3.09 (1.90-5.02)** |
| **Ever watched pornographic film** |  |  |  |  |
| No | 318 | 84 | 1 | 1 |
| Yes | 69 | 51 | 2.80(1.81-4.32)** | 1.55(0.93-2.58) |
| **Attend night clubs** |  |  |  |  |
| No | 361 | 102 | 1 | 1 |
| Yes | 26 | 33 | 4.49(2.57-7.86)** | 2.56(1.35-4.86)* |
| **Alcohol drinking frequency** |  |  |  |  |
| Never | 338 | 89 | 1 | 1 |
| Sometimes | 49 | 46 | 3.57(2.24-5.68)** | 1.90(1.10-3.29)* |
| **Parental neglect** |  |  |  |  |
| Yes | 237 | 48 | 2.86(1.91-4.30)** | 2.10(1.35-3.29)** |
| No | 150 | 87 | 1 | 1 |
| **HIV/AIDS Knowledge** |  |  |  |  |
| Good | 190 | 44 | 1 | 1 |
| Poor | 197 | 91 | 2.00(1.32-3.01) | 1.76(1.12-2.77)* |

\*= P value < 0.05; \*\*= P value < 0.01

Respondents whose close friend experienced sex were threefold-fold (adjusted odds ratio [AOR] = 3.09; 95% CI (confidence interval): 1.90-5.02) more likely to have risky sexual behavior than respondents whose close friend did not experience sex. In contrast to students who do not attend night clubs, those students who attend night clubs are 2.56 times (AOR=2.56; 95% CI: 1.35-4.86) more prone to risky sexual behavior. A respondent who sometimes drinks alcohol 1.90 times (AOR=1.90; 95% CI: 1.10-3.29) tends to have risky sexual behavior more than a respondent who never drinks alcohol. The odds of having risky sexual behavior among students with parental neglect is twice (AOR=2.10; 95% CI: 1.35-3.29) higher than that among students with no parental neglect. Participants with poor HIV/AIDS knowledge were 1.76 times (AOR=1.76; 95% CI: 1.12-2.77) more likely to have risky sexual behavior than participants with good HIV/AIDS knowledge.

## Discussion

Given the high burden of HIV/AIDS infection in the South Omo zone and limited scientific information on the prevalence of RSB and its determinants among secondary school students in the study area, this study was deemed necessary.

This study showed that one-fourth (25.9%) of secondary school students in the South Omo zone had experienced risky sexual behavior. The findings of this study are higher than those of a study conducted in Aksum town (17.2%) (24) and in Jiga town (14.7%) (25). The observed variation may be explained by the difference in methodologies and socioeconomic status. Moreover, a possible explanation for the discrepancy may be the difference in the time of study. Hence, students in the current study are more likely to watch pornographic movies and attend night clubs due to globalization and technological advances.

On the other hand, the magnitude of RSB in our study is less than the findings from studies in Addis Ababa (26.7%) (26), Kigali (41%) (27) and Bangkok (69.5%) (28). The possible reason for the difference may be the difference in the study setting in that the current study involved a more rural setting where there is limited technology to access events such as pornography.

Students whose close friends experienced sexual intercourse had increased odds of engaging in risky sexual behavior than their counterparts in this study setting. This is in line with past studies conducted in Wolaita Sodo town (29) and Enemay district (18). This could be due to peer pressure that could affect the behaviors of youths in a negative way, as shown by the study in Kigali (27).

The current study also revealed that in contrast to students who do not attend night clubs, those students who attend night clubs are more than double as prone to practice risky sexual behavior. This finding is supported by a previous study in Ethiopia (30). This can be explained by the fact that students who attend night clubs frequently may have alcohol or smoking addiction, which can lead them to lose control over their sexual behaviors that could lead to risky sexual behavior. In harmony with prior studies in Nigeria (31) and in Ethiopia (32), this study discovered that a higher frequency of alcohol consumption increases the chance of having RSB. This could be related to the fact that consuming alcohol and drugs would increase risk-taking behavior, impair one’s judgment on the risk of unprotected sexual behaviors and impair sexual health decision-making. Furthermore, drinking alcohol prior to sexual intercourse was believed to provide a socially acceptable excuse for nonuse of condoms.

Consistent with a study carried out in Aksum town (19), this study showed that parental neglect increases the chance of experiencing risky sexual behavior among secondary students. Perhaps this is due to a lack of parental monitoring and discussion with parents on sexual matters that will lead to the practice of risky sexual behavior due to being free from parental supervision and poor decision making on sexual and reproductive matters, as revealed in various studies (27, 29, 33).

Having good HIV/AIDS-related knowledge among secondary students was associated with a lower risky sexual behavior experience in the current study area. This association is in agreement with a study performed in Arsi Negele (17). This could be attributed to students who have knowledge of the transmission of HIV/AIDS through unprotected sexual intercourse and having multiple sexual partners refraining from participating in such risky sexual behaviors.

Due to the employment of a cross-sectional study design, this study suffers from the chicken or egg dilemma regarding the temporal relation between the outcome variable and independent variables. This study also has a limitation of not including all youths in the study area.

## Conclusions

One-fourth of the participants in this study engaged in risky sexual behavior, and frequent alcohol consumption, parental neglect, having sexually active close friends, poor knowledge of HIV/AIDS, and nightclub attendance were factors that were significantly positively associated with risky sexual behavior.

The results of this study highlight the importance of life skill training to tackle peer pressure, working to ease alcohol consumption and night club attendance to reduce the practice of risky sexual behavior among secondary school students in the South Omo Zone. Due emphasis should also be given to integrating adolescent health into health care service delivery by health extension workers and health workers while strengthening sexual and reproductive health services to youths and adolescents to enhance HIV/AIDS-related knowledge and informed decision making. Parents must be well informed about the developmental requirements of their teenager and promote open communication regarding sexual and reproductive health within the family. Finally, more studies should be conducted considering all youths and using prospective cohort or experimental study designs to explore the temporal relationship between various factors and RSB.

## Declarations

## Data Availability

The datasets used and/or analyzed during the current study are available from the corresponding author on reasonable request.

## List of abbreviations

AIDS: Acquired Immunodeficiency Disease
AOR: AAdjusted Odds Ratio
CI: Confidence Interval
HIV: Human Immunodeficiency Virus
RSB: Risky Sexual Behavior
SNNPR: Southern Nations, Nationalities and People’s Region

## Acknowledgments

The authors are grateful to Jinka University for financial support and ethical clearance to conduct this work. Our acknowledgment also goes to thank the South Omo zonal health department for giving us permission to carry out the research and providing some relevant information. We extend our heartfelt appreciation to the data collectors, the supervisors and study participants without whom this work had been accomplished.

## Authors’ contributions

GAT wrote the study proposal, including its methods and work plan, in addition to analyzing the data and writing the results. GAT, EWW, MDA and BAA supervised the data collection and wrote the discussion and conclusion. All authors read and approved the final manuscript.

## Ethics approval and consent to participate

Ethical Review Committee of Jinka University gave ethical clearance for this study (reference number: JKU/RCE/ERC/041/15). Confidentiality was guaranteed by excluding personal identifiers, and informed consent was obtained from the respondents. Respondents were allowed to not participate in the study or withdraw at any time during the study, upon their request.

## Consent for publication

Not applicable.

